# Life-course socioeconomic factors are associated with markers of epigenetic aging in a population-based study

**DOI:** 10.1101/2022.06.16.22276489

**Authors:** Dusan Petrovic, Cristian Carmeli, José Luis Sandoval, Barbara Bodinier, Marc Chadeau-Hyam, Stephanie Schrempft, Georg Ehret, Nasser Abdalla Dhayat, Belén Ponte, Menno Pruijm, Emmanouil Dermitzakis, Paolo Vineis, Sémira Gonseth-Nusslé, Idris Guessous, Cathal McCrory, Murielle Bochud, Silvia Stringhini

## Abstract

Adverse socioeconomic circumstances negatively affect the functioning of biological systems, but the underlying mechanisms remain only partially understood. Here, we explore the associations between life-course socioeconomic factors and four markers of epigenetic aging in a population-based setting.

We used data from a population-based study conducted in Switzerland (SKIPOGH) to assess the association between childhood, adulthood, and life-course socioeconomic indicators, and blood-derived markers of epigenetic aging (Levine’s, DunedinPoAm38, GrimAge epigenetic age acceleration (EAA) and the mortality risk score (MS)). We used mixed regression to explore the associations between socioeconomic indicators and markers of epigenetic aging independently, and counterfactual mediation to investigate the mechanisms underlying the life-course socioeconomic gradient in epigenetic aging.

Individuals reporting a low father’s occupation, adverse financial conditions in childhood, a low income, having financial difficulties, or experiencing unfavorable socioeconomic trajectories were epigenetically older than their more advantaged counterparts. Specifically, this corresponded to an average increase of 1.0-1.5 years for Levine’s epigenetic age when compared to chronological age, 1.1-1.5 additional years for GrimAge, 5%-8% higher DunedinPoAm38 EAA, and 2%-5% higher MS score. By exploring the life-course mechanisms underlying the socioeconomic gradient in epigenetic aging, we found that both childhood and adulthood socioeconomic factors contributed to epigenetic aging, and that detrimental lifestyle factors mediated the relation between socioeconomic circumstances in adulthood and EAA.

Our study provides novel empirical evidence for a “sensitive-period” life-course model, whereby adverse socioeconomic circumstances in childhood *and* adulthood negatively affected epigenetic aging. Counterfactual mediation analyses further showed that the effect of socioeconomic factors in adulthood operated through detrimental lifestyle factors, whereas associations involving early-life socioeconomic factors were less clear.

## 1 Introduction

The disruption of major physiological systems constitutes a key step in the biological “embedding” of the social environment, and plays a central role in the occurrence of disease and premature mortality [1, 2]. These processes include aberrant inflammation, hormonal dysregulation, impaired neural function, poor metabolic control, or allostatic load, with socially disadvantaged individuals consistently displaying adverse physiological patterns in a dose-response manner [1, 3-7]. Furthermore, the ever-growing evidence for a contribution of socially-driven epigenetic modifications has been the object of particular attention in recent years [1, 8, 9].

Epigenetic modifications may result from changes in DNA methylation, which refer to the addition or removal of methyl groups to Cytosine-phospho-Guanine (CpG) dinucleotides across the genome [10]. DNA methylation changes occur naturally during development and senescence, but may also result from environmental exposures, including lifestyle factors, pollutants, or socioeconomic adversity, eventually contributing to an increased disease risk [9, 11-15]. DNA methylation may therefore provide a candidate mechanism for the biological embedding of social exposures [15-17]. Recent years witnessed the development of the so-called *epigenetic clocks*, which are based on specific sets of age-dependent CpGs and allow for determination of whether an individual is experiencing *epigenetic age acceleration* (EAA), which has been related to adverse socioeconomic conditions across the life-course, but also to increased disease and mortality risk [9, 16, 18-21]. First generation epigenetic clocks were designed to be highly accurate predictors of chronological age, and measure changes in DNA methylation shared between individuals. More recently, second generation epigenetic clocks have been designed to gauge inter-individual variability in aging and were trained using health-related biomarkers in addition to age [15, 16, 22, 23].

In this study, we sought to characterize the associations between socioeconomic factors and markers of epigenetic aging using data from a Swiss population-based study.

Focusing on health-related second generation epigenetic markers, we first examine the associations between multiple socioeconomic indicators in childhood and adulthood, and Levine, DunedinPoAm38, and GrimAge clocks, as well as the mortality risk score (MS). Second, we explore the life-course models and the contribution of intermediate factors through which socioeconomic factors operate to affect epigenetic aging. Using counterfactual mediation, we investigate the mutual relations involving childhood and adulthood socioeconomic indicators and epigenetic aging in the light of three life-course scenarios: the “critical period” model, which postulates that there are time windows throughout life when the body is particularly sensitive to adverse exposures (i.e. *in utero* development, the first year of life, adolescence, etc.); the “chains of risk” model, implying that a sequence of linked exposures eventually affect one’s disease risk; and the “social mobility” model, whereby the direction of socioeconomic mobility across the life-course (i.e. upward/downward) has an impact on subsequent biological processes and health events [24, 25]. Finally, we examine the contribution of lifestyle exposures (smoking, alcohol consumption, sedentary behavior, Body Mass Index (BMI)), which have been previously identified as important mediators of the socioeconomic gradient in health-related outcomes and epigenetic age acceleration [26-28].

## 2 Material and methods

### 2.1 Study population

We used data from the Swiss Kidney Project on Genes in Hypertension (SKIPOGH), a family- and population-based cohort investigating the genetic and environmental determinants of health-related outcomes in the Swiss population. Study participants were recruited in the city of Lausanne and the cantons of Geneva and Bern between 2009 and 2013 (SKIPOGH 1, baseline visit), and came for a follow-up visit three years later (SKIPOGH 2, present sample) [4]. Study inclusion criteria were: 1. Written informed consent; 2. 18 years of age; 3. Caucasian origin; 4. At least one first-degree family member willing to participate to the study. Women who reported being pregnant were excluded from the study. At both visits, included participants attended a medical examination after an overnight fast, provided blood and urine samples, and completed a self-administered questionnaire inquiring about their living standards, socioeconomic and financial circumstances in childhood and adulthood, lifestyle factors, and medical history. All participants provided written informed consent.

### 2.2 Socioeconomic indicators

We used nine socioeconomic indicators in early-life and adulthood as the main exposure variables. Socioeconomic indicators in early-life focused on participant’s childhood, inquiring about father’s occupational position, financial and material conditions the participants enjoyed in childhood, and father’s and mother’s highest achieved education (**Appendix A**). Socioeconomic indicators in adulthood included participant’s own education, last known occupational position, monthly household income, reporting financial difficulties, and forgoing healthcare due to economic reasons. All nine indicators were self-reported following predefined categories, and subsequently recoded into three (“High”/“No financial difficulties” (Reference group - most favorable), “Middle”/“Average financial difficulties”, and “Low”/“Important financial difficulties”) or two groups (“Not forgoing healthcare” (Reference group - most favorable), “Forgoing healthcare”), as described in **Appendix A**. We further generated a proxy for life-course inter-generational social mobility based on father’s occupational position and participant’s last known occupational position. Five socioeconomic trajectories were possible: “Stable high” (Reference group: high father’s occupation and high own occupation), “Upward mobile” (low father’s occupation and middle/high own occupation, or middle father’s occupation and high own occupation), “Stable middle” (middle father’s occupation, middle own occupation), “Downward mobile” (high father’s occupation and middle/low own occupation, or middle father’s occupation and low own occupation), “Stable low” (low father’s occupation and low own occupation).

### 2.3 CpG DNA methylation measurement and data pre-processing

Epigenome-wide DNA methylation from white blood cells was measured in 242 SKIPOGH participants using the Infinium HumanMethylation450 BeadChip microarray of Illumina (HM450), measuring the methylation status at 485,512 CpG sites. For a different set of 442 SKIPOGH participants, epigenome-wide DNA methylation was measured using a more recent Infinium MethylationEPIC v1.0 microarray (EPIC), including >90% of the CpG sites from the HM450 and an additional 413,743 CpGs (865,859 CpGs in total) [29]. For both arrays, CpG methylation data were summarized as β coefficients representing a ratio of the average signal for methylated CpG sites to the sum of methylated *and* unmethylated sites. To control for the noise introduced by technical factors (methylation array type, array position, and plate level), the CPACOR procedure was applied, yielding 30 principal components to be used as fixed-effect covariates [30]. Missing data was imputed using the nearest averaging multiple imputation method [31]. The data preprocessing procedures eventually yielded 452,453 CpG sites available for analyses.

### 2.4 Epigenetic age acceleration and mortality risk score

Using blood-derived DNA methylation, we computed four markers of epigenetic aging: Levine’s epigenetic clock (DNAmPhenoAge), Dunedin Pace of Aging Methylation (DunedinPoAm38), GrimAge epigenetic clock, and the mortality risk score (MS) [16, 20, 22, 28]. The first three epigenetic clocks (Levine, DunedinPoAm38, GrimAge) display a continuous distribution and correlate positively with chronological age (**Supplementary Figure 1**), whilst the MS score displays a discrete distribution, ranging from 0 (lowest risk – no CpG in the risk quartile) to 10 (highest risk – all ten CpGs in the risk quartile). We subsequently computed *epigenetic age acceleration* (EAA) for Levine, DunedinPoAm38, and GrimAge epigenetic clocks, calculated as the residuals from the regression of epigenetic clocks (response) on chronological age, CPACOR principal components, and chip type (predictors), with positive EAA values representing an accelerated or mitigated epigenetic aging [9]. Hereinafter, we will refer to EAA and MS (main outcome variables) as *markers of epigenetic aging*.

We chose to include these markers as they have been specifically elaborated using CpGs related to chronological age *and* multiple health-related biomarkers, and may possibly reflect health deterioration as a result of external stressors, including socioeconomic adversity and poor lifestyle factors [9, 19, 32]. Further, all four markers have been developed using DNA methylation profiles derived from blood, which is routinely sampled in population-based studies [17, 18, 32]. Additional details on the calculation of the included markers are provided in **Appendix B**.

### 2.5 Statistical analyses

We applied mixed regression models to estimate the *total effect* of life-course socioeconomic factors (main exposure variables) on markers of epigenetic aging (main outcome variables). We used childhood, adulthood, and life-course socioeconomic factors as independent, *categorical* exposure variables. To further explore for potential dose-response effects driven by socioeconomic indicators and to implement counterfactual mediation analyses, we generated *continuous* summary measures for each socioeconomic indicator (lowest vs. highest, (LH)), hypothesizing a linear effect on markers of epigenetic aging [33]. We implemented mixed *linear* regression when using Levine, DunedinPoAm38, and GrimAge EAA as the outcome variables (continuous), and *Poisson* regression when using the MS score (discrete variable). All regression models were adjusted for age, sex, study center, seasonality of blood sampling, and familial structure (random-effect variable). For analyses using adulthood socioeconomic indicators as the main exposure variable, we accounted for the potential confounding effect of *childhood* socioeconomic indicators by including them as additional covariates [2].

We imputed missing data (socioeconomic indicators in childhood, adulthood and life-course trajectories) through multiple imputation by chained equations (MICE) by hypothesizing missingness at random (5 imputed data sets, 50 iterations). All statistical analyses were carried out using the R statistical software and relevant CRAN and Bioconductor packages (R Foundation for Statistical Computing, Vienna, Austria).

### 2.6 Counterfactual mediation analyses and life-course models

To investigate the intermediate mechanisms through which life-course socioeconomic indicators affect epigenetic aging, we implemented two *counterfactual mediation models* in order to assess the contribution of intermediate factors (mediators) located on the causal pathway between the main exposure (socioeconomic indicators) and the main outcome (markers of epigenetic aging) variables. Briefly, counterfactual mediation analysis allows for decomposing the *total effect* estimated in the main analyses into a *Natural Direct Effect* (NDE), which represents the relation between the exposure and the outcome through pathways which *do not involve* the mediators, and a *Natural Indirect Effect* (NIE), which represents the relation between the exposure and outcome via pathways *involving* the mediators. The contribution of the mediators to the association between the exposure and the outcome is estimated via the *Proportion Mediated* parameter (PM), defined as the ratio between NIE and the *Marginal Total Effect* (MTE=NDE+NIE) [34].

In the first mediation model, we sought to determine whether the association between socioeconomic factors *in childhood* (exposure: lowest vs. highest) and markers of epigenetic aging (outcome), operates through socioeconomic and lifestyle factors *in adulthood* (mediators: adulthood SE indicators, smoking, hazardous alcohol intake, sedentary behavior, BMI (**Appendix C**)) (**Figure 1A**). We included these mediators as previous research has reported that early-life socioeconomic circumstances act as determinants of socioeconomic status in adulthood, which in turn affect physiological and aging processes through lifestyle factors [2, 25, 26]. From the life-course perspective, the implementation of this model would allow disentangling whether childhood socioeconomic factors affect markers of epigenetic aging according to the “critical period” model, whereby adverse exposures leave a permanent imprint on physiological processes during periods when the body is particularly sensitive (i.e. *in utero* development, first year of life, adolescence), or according to “pathway models” (i.e. “chains of risk”), implying that childhood exposures affect physiological processes via subsequent exposures in adulthood [24, 25]. In this particular framework, a null indirect effect (NIE) would tend to support a “critical period” model, whereas the presence of a non-null indirect effect would support “chains of risk” models.

**Figure 1:**
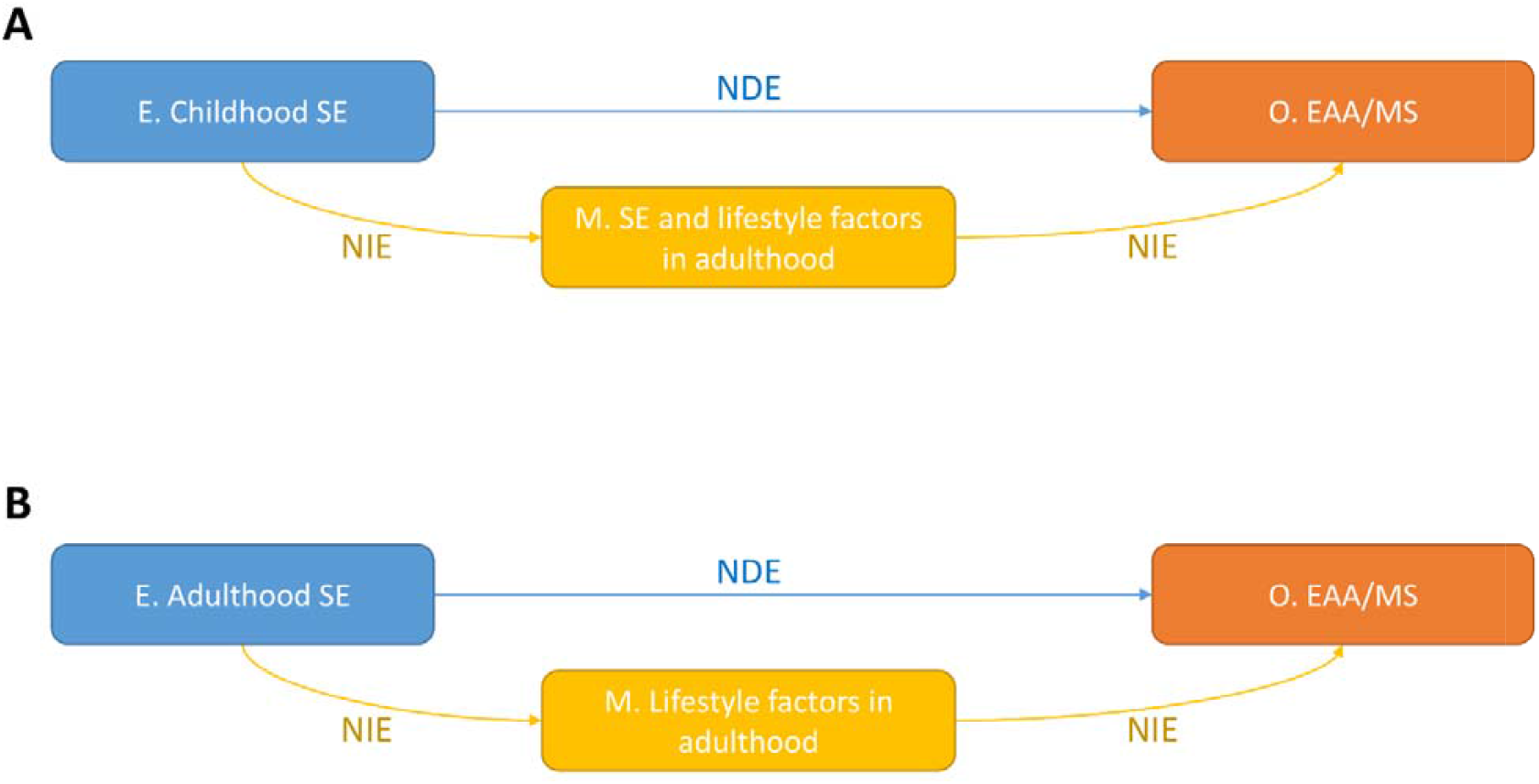
Directed Acyclic Graphs (DAG) representing the causal structure of the counterfactual mediation models implemented in this study. (A) DAG representing the association between childhood socioeconomic factors (exposure) and markers of epigenetic aging (outcome: EAA/MS), using adulthood socioeconomic factors and lifestyle factors (smoking, alcohol intake, physical inactivity, and BMI) as *en-bloc* mediators. (B) DAG representing the association between socioeconomic factors in adulthood and markers of epigenetic aging using lifestyle factors as en-bloc mediators. E, Exposure; M, Mediators, O, Outcome. NDE, Natural Direct Effect representing the relation between the exposure and outcome via pathways which do not involve the mediators. NIE, Natural Indirect Effect representing the relation between the exposure and outcome via the mediators (en-bloc). MTE, Marginal Total Effect (not depicted) computed as the sum of NDE and NIE. Confounding variables (age, sex, study center, seasonality of blood sampling, and familial structure) are not represented for the sake of simplicity.

In the second mediation model, we aimed to specifically assess the contribution of self-reported lifestyle factors (**Appendix C**: smoking, hazardous alcohol intake, sedentary behavior, and BMI *in adulthood*) to the association between socioeconomic indicators *in adulthood* and markers of epigenetic aging (**Figure 1B**). We chose to include these mediators as previous research has consistently shown that socioeconomic factors in adulthood act as direct determinants of lifestyle factors, which in turn affect multiple biological processes, including epigenetic aging [9, 11, 18, 26].

For both counterfactual models, we adopted a “regression-based” counterfactual mediation analysis which incorporates interaction terms between the exposure and the mediators, and evaluated a joint mediating effect by the mediators (*en-bloc mediators*) [9, 35, 36]. Missing values were imputed using multiple imputation chained equations (in-built R package method) [36]. Standard errors for all mediation parameters (MTE, NDE, NIE, PM) were estimated through percentiles from 1000 bootstrap draws with replacement [36].

### 2.7 Supplementary analyses

Considering that white blood cells (WBC) composition may influence DNA methylation patterns, we repeated the main analyses by computing *intrinsic* epigenetic age acceleration and MS score, which calculates EAA residuals by including Houseman-estimated WBC composition as additional predictors, or by directly including WBC composition as additional covariates in the model (MS score) [37, 38].

Further, as epigenetic clocks may not be optimally calibrated for older individuals and since epigenetic age tends to increase at a different pace across age groups, we repeated the main regression models by stratifying the associations at 65 years of age [39, 40].

## 3 Results

### 3.1 Characteristics of study population

The mean age was 52.6 years, with 52% of the study population being female, as reported in **Table 1**. Approximately one quarter of included participants reported a high father’s occupational position in childhood (24%), high financial conditions in childhood (26%), and a high father’s education level (25%), whereas 13% of participants declared a high mother’s education in childhood. In adulthood, 39% reported having achieved high education, 21% reported a high occupational position, and 38% were in the high household income group. Related to everyday financial difficulties and their consequences, 67% declared experiencing no financial difficulties in everyday life, and 90% reported not having forgone healthcare during the previous year due to economic reasons. 25% of included participants were current smokers, 5% declared having a hazardous alcohol intake, 40% reported being sedentary, whereas the average BMI was 25.6 kg/m^2^. Finally, we observed that the GrimAge epigenetic clock displayed a mild deviation from the chronological age (54.6 years), while the average values of DunedinPoAm38 and MS score were 0.9 and 2, respectively.

**Table 1:**
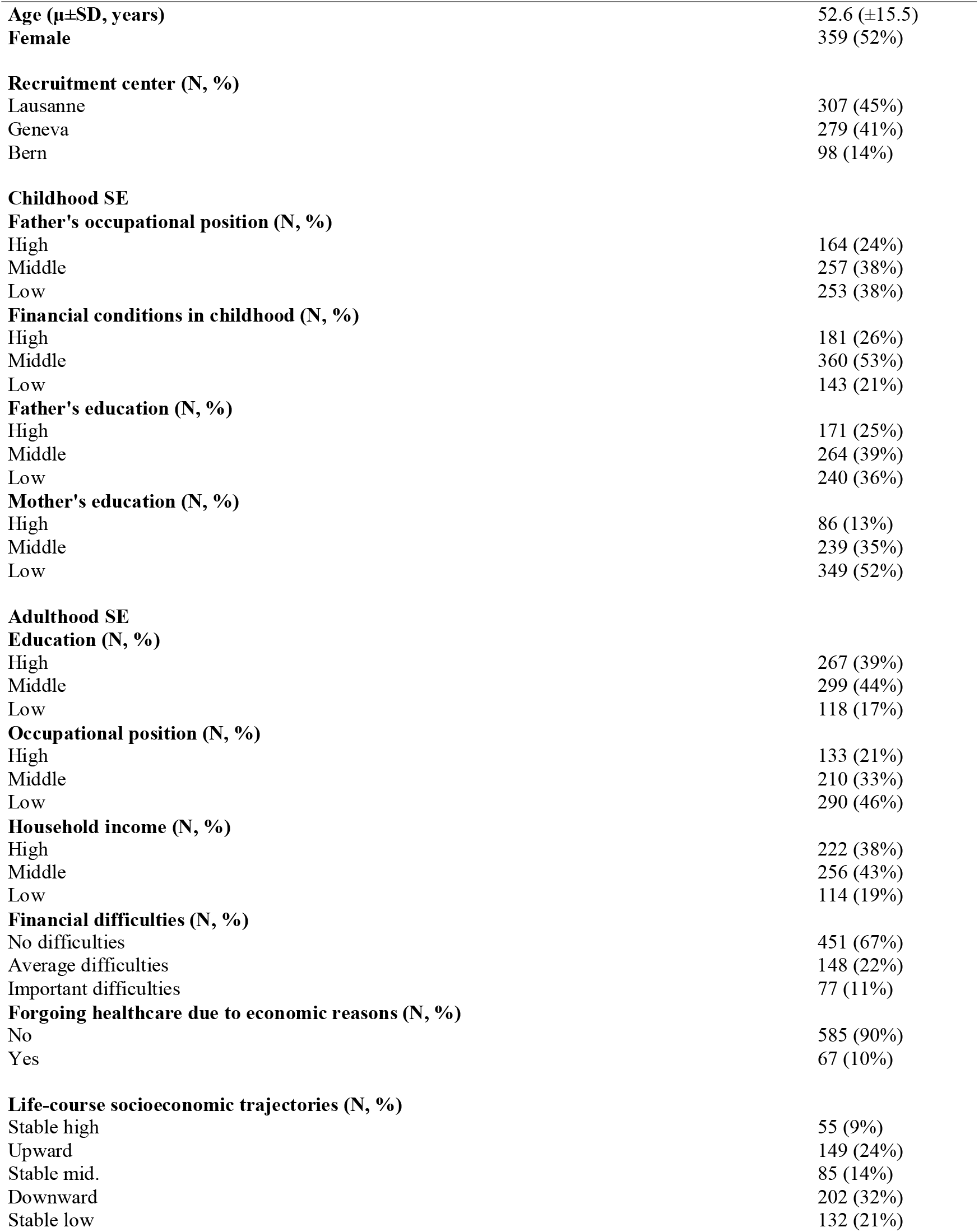
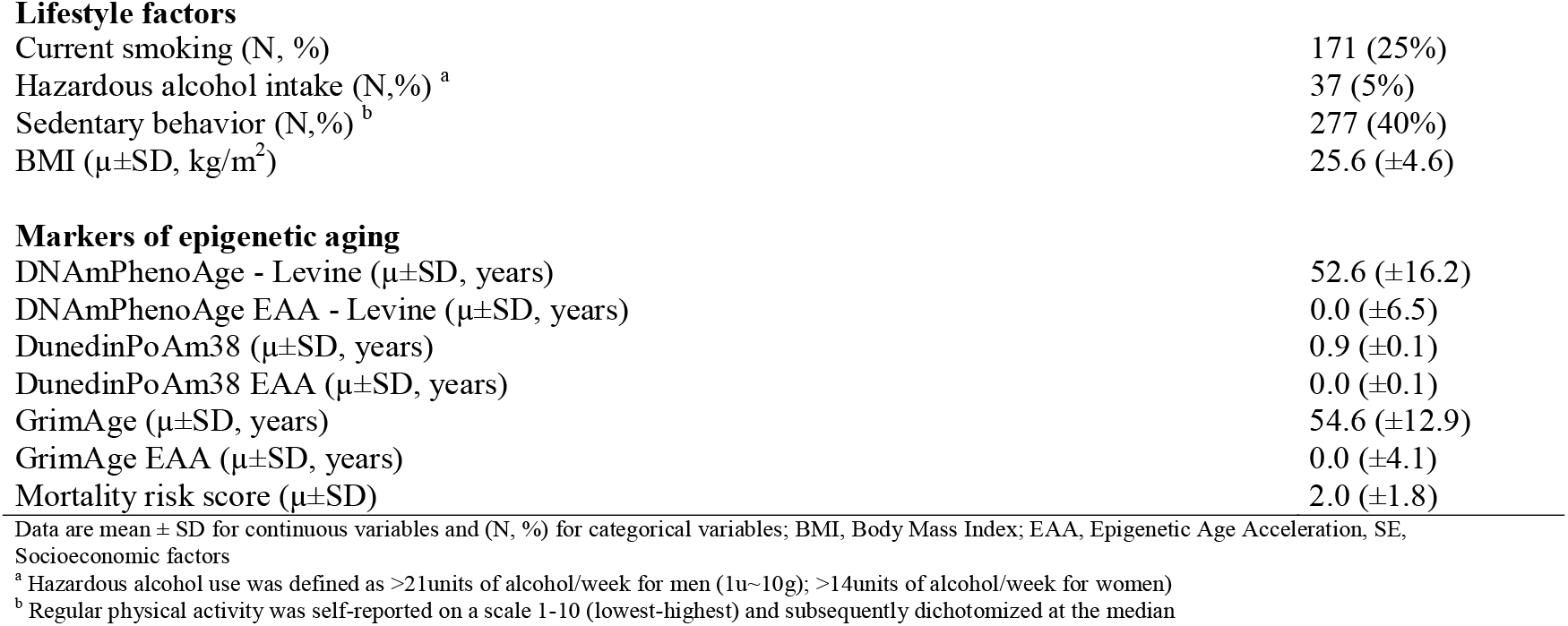
General characteristics of the study population

|  |  |
| --- | --- |
| **Age ($\mu \pm SD$, years)** | 52.6 ($\pm 15.5$) |
| **Female** | 359 (52%) |
| **Recruitment center (N, %)** |  |
| Lausanne | 307 (45%) |
| Geneva | 279 (41%) |
| Bern | 98 (14%) |
| **Childhood SE** |  |
| **Father's occupational position (N, %)** |  |
| High | 164 (24%) |
| Middle | 257 (38%) |
| Low | 253 (38%) |
| **Financial conditions in childhood (N, %)** |  |
| High | 181 (26%) |
| Middle | 360 (53%) |
| Low | 143 (21%) |
| **Father's education (N, %)** |  |
| High | 171 (25%) |
| Middle | 264 (39%) |
| Low | 240 (36%) |
| **Mother's education (N, %)** |  |
| High | 86 (13%) |
| Middle | 239 (35%) |
| Low | 349 (52%) |
| **Adulthood SE** |  |
| **Education (N, %)** |  |
| High | 267 (39%) |
| Middle | 299 (44%) |
| Low | 118 (17%) |
| **Occupational position (N, %)** |  |
| High | 133 (21%) |
| Middle | 210 (33%) |
| Low | 290 (46%) |
| **Household income (N, %)** |  |
| High | 222 (38%) |
| Middle | 256 (43%) |
| Low | 114 (19%) |
| **Financial difficulties (N, %)** |  |
| No difficulties | 451 (67%) |
| Average difficulties | 148 (22%) |
| Important difficulties | 77 (11%) |
| **Forgoing healthcare due to economic reasons (N, %)** |  |
| No | 585 (90%) |
| Yes | 67 (10%) |
| **Life-course socioeconomic trajectories (N, %)** |  |
| Stable high | 55 (9%) |
| Upward | 149 (24%) |
| Stable mid. | 85 (14%) |
| Downward | 202 (32%) |
| Stable low | 132 (21%) |
| **Lifestyle factors** |  |
| Current smoking (N, %) | 171 (25%) |
| Hazardous alcohol intake (N, %) <sup>a</sup> | 37 (5%) |
| Sedentary behavior (N, %) <sup>b</sup> | 277 (40%) |
| BMI ($\mu \pm$SD, kg/m<sup>2</sup>) | 25.6 (±4.6) |
| **Markers of epigenetic aging** |  |
| DNAmPhenoAge - Levine ($\mu \pm$SD, years) | 52.6 (±16.2) |
| DNAmPhenoAge EAA - Levine ($\mu \pm$SD, years) | 0.0 (±6.5) |
| DunedinPoAm38 ($\mu \pm$SD, years) | 0.9 (±0.1) |
| DunedinPoAm38 EAA ($\mu \pm$SD, years) | 0.0 (±0.1) |
| GrimAge ($\mu \pm$SD, years) | 54.6 (±12.9) |
| GrimAge EAA ($\mu \pm$SD, years) | 0.0 (±4.1) |
| Mortality risk score ($\mu \pm$SD) | 2.0 (±1.8) |

Data are mean ± SD for continuous variables and (N, %) for categorical variables; BMI, Body Mass Index; EAA, Epigenetic Age Acceleration, SE, Socioeconomic factors
<sup>a</sup> Hazardous alcohol use was defined as >21units of alcohol/week for men (1u~10g); >14units of alcohol/week for women)
<sup>b</sup> Regular physical activity was self-reported on a scale 1-10 (lowest-highest) and subsequently dichotomized at the median

### 3.2 Socioeconomic factors and markers of epigenetic aging

In **Figure 2**, we describe the associations between socioeconomic factors and markers of epigenetic aging. Reporting adverse financial conditions in childhood, important financial difficulties in adulthood, and experiencing less favorable socioeconomic trajectories across the life-course (all but stable high) were associated with increased Levine’s epigenetic age acceleration (**Panel A** (*categories*): Adverse financial conditions in childhood: +1.49 years 95%CI[-0.04;3.04], Important financial difficulties in adulthood: 1.52 years 95%CI[0.05;2.98]; Upward socioeconomic trajectory: 2.13 years 95%CI[0.37;3.90], Stable low socioeconomic trajectory: 2.23 years 95%CI[0.41;4.05]). Further examining “Lowest vs. Highest” regression gradients (β_LH_), we found no dose-response relationship between *linearized* socioeconomic indicators and Levine EAA. We observed similar associations between adulthood and socioeconomic indicators in adulthood and across the life-course (household income, financial difficulties, forgoing healthcare due to economic reasons, stable low socioeconomic trajectory) with DunedinPoAm38 and GrimAge EAA markers (**Panels B-C**), with less favorable socioeconomic factors being associated with increased EAA, whereas reporting low financial conditions in childhood was associated with increased GrimAge EAA (1.20 years 95%CI[0.17;2.23]). Using *linearized* socioeconomic indicators, we further observed consistent dose-response effects between socioeconomic factors in adulthood and DunedinPoAm38 and GrimAge EAA, as well as between adverse financial conditions in childhood and increased GrimAge EAA. Finally, we found that reporting low financial conditions in childhood, having a mid-level education, a low occupational position, a low household income, and experiencing less favorable socioeconomic trajectories across the life-course was consistently associated with an increased MS score (**Panel D** (*categories*): Low financial conditions in childhood: 0.19 (2.4% higher MS score) 95%CI[0.00;0.39]; Low occupation: 0.20 95%CI[0.03;0.37], Low household income: 0.37 95%CI[0.20;0.54], Stable low trajectory: 0.29 95%CI[0.03;0.54]). Furthermore, examining regression coefficients for *linearized* socioeconomic indicators, we found that financial conditions in childhood and household income in adulthood displayed a consistent dose-response effect with the MS score.

**Figure 2:**
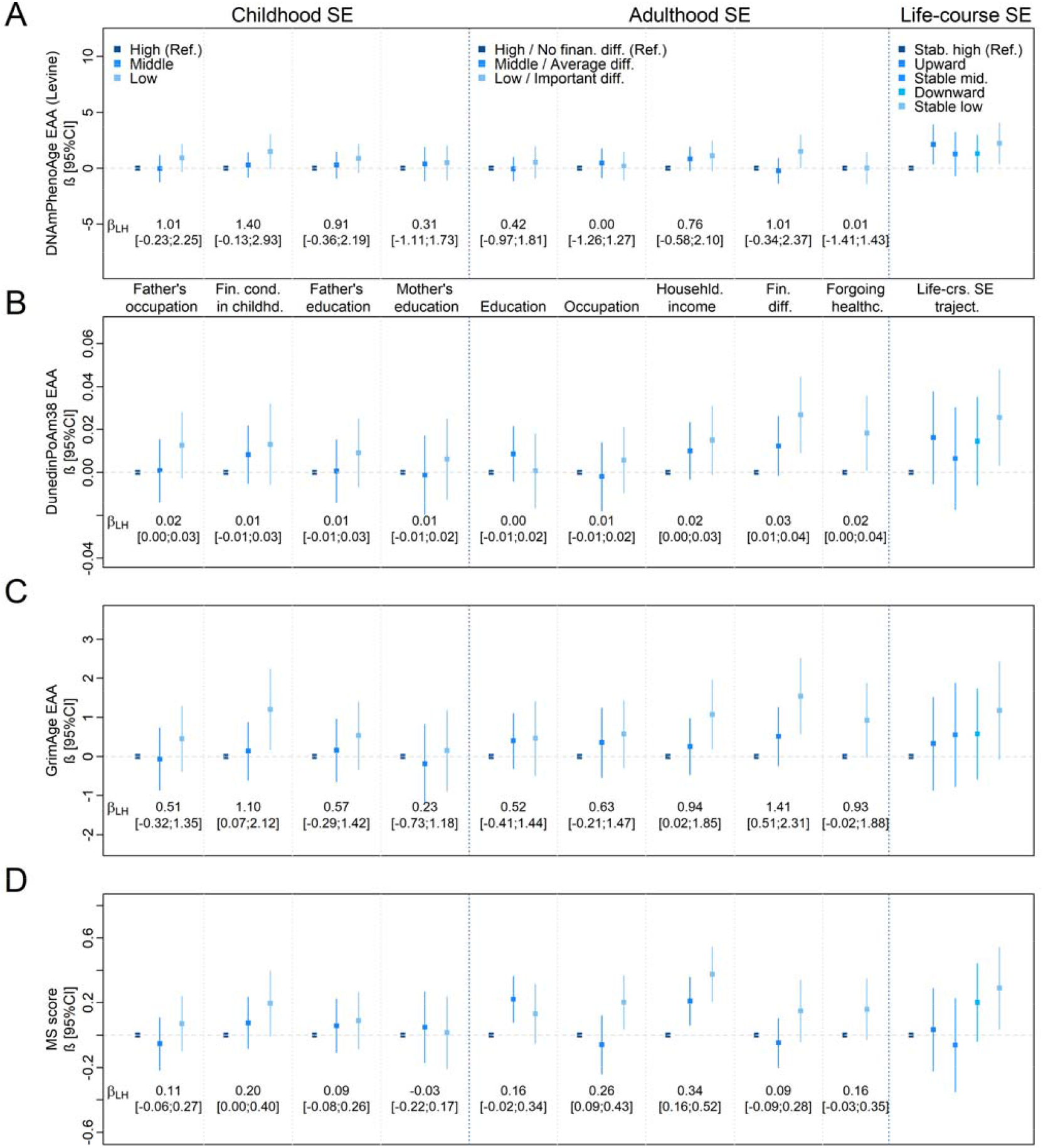
Regression coefficients for the associations between life-course socioeconomic factors and markers of epigenetic aging EAA, Epigenetic age acceleration; Fin. cond., financial conditions; Fin. diff., Financial difficulties; Life-crs. SE traject, Life-course socioeconomic trajectories; LH, lowest versus highest; SE, socioeconomic indicators Forgoing healthcare due to economic reasons was used as a two-categorical predictor: Not forgoing healthcare (Ref.); Forgoing healthcare Regression models for the association between life-course socioeconomic indicators (categorical predictor) and second generation epigenetic age acceleration (ABC, continuous outcome) and mortality risk score (D, discrete outcome), adjusted for sex, age, center, season, familial structure (random effect). Associations involving socioeconomic indicators in adulthood were additionally adjusted for early-life socioeconomic indicators β_LH_: Regression coefficient for socioeconomic indicators used as *continuous (linearized)* predictors (Lowest vs. Highest) Missing data was imputed for socioeconomic indicators (childhood, adulthood, life-course) using multivariate imputation by chained equations and by hypothesizing missingness at random (5 imputed datasets, 50 iterations)

### 3.3 Counterfactual mediation analyses

In **Table 2**, we present the counterfactual mediation estimates for the associations between socioeconomic indicators in childhood (exposure), socioeconomic and lifestyle factors in adulthood (mediators), and markers of epigenetic aging (outcome).We found no *total* (MTE), *direct* (NDE), *indirect* (NIE), nor *mediating effect* (PM) by socioeconomic and lifestyle factors in adulthood, except for an association between financial conditions in childhood and GrimAge EAA (MTE: 1.24 95%CI[0.09;2.47]).

**Table 2:** Counterfactual mediation estimates for the associations between childhood socioeconomic indicators, socioeconomic and lifestyle factors in adulthood, and markers of epigenetic aging (DAG: Figure 1A)

| **DNAmpHenoAge acceleration (Levine)** | **MTE** | **NDE** | **NIE** | **PM** |
| --- | --- | --- | --- | --- |
| Father's occupational position | 1.43[-1.15;3.25] | 0.63[-2.15;2.64] | 0.08[-0.71;0.88] | 0.06[-3.43;2.14] |
| Financial conditions in childhood | 1.55[-0.16;3.42] | 1.31[-0.50;3.30] | 0.25[-0.53;1.16] | 0.16[-1.17;1.28] |
| Father's education | 2.09[-1.66;3.67] | 1.48[-2.43;3.26] | -0.04[-0.70;0.67] | -0.02[-1.94;3.04] |
| Mother's education | -0.30[-2.45;3.25] | -1.17[-3.73;2.42] | 0.06[-0.45;0.51] | -0.20[-2.22;2.78] |
| **DunedinPoAm38 age acceleration** | **MTE** | **NDE** | **NIE** | **PM** |
| Father's occupational position | 0.02[-0.01;0.03] | 0.01[-0.01;0.03] | 0[-0.02;0.01] | -0.1[-3.72;3.30] |
| Financial conditions in childhood | 0.02[-0.01;0.04] | 0.01[-0.02;0.03] | 0.01[-0.01;0.02] | 0.36[-3.49;2.41] |
| Father's education | 0.00[-0.03;0.04] | 0.00[-0.04;0.04] | 0.00[-0.01;0.01] | 1.20[-4.67;4.59] |
| Mother's education | -0.01[-0.03;0.04] | -0.02[-0.05;0.03] | 0.01[0.00;0.02] | -1.53[-5.4;10.10] |
| **GrimAge acceleration** | **MTE** | **NDE** | **NIE** | **PM** |
| Father's occupational position | 0.57[-0.49;1.62] | 0.27[-0.75;1.35] | -0.06[-0.79;0.54] | -0.1[-4.62;5.85] |
| Financial conditions in childhood | **1.24[0.09;2.47]** | 0.86[-0.33;2.13] | 0.35[-0.45;1.19] | 0.29[-0.91;1.65] |
| Father's education | 1.57[-1.29;2.15] | 1.42[-1.23;2.02] | 0.19[-0.66;0.79] | 0.12[-5.88;4.46] |
| Mother's education | -0.42[-2.65;2.94] | -0.97[-3.26;2.51] | 0.48[-0.09;1.03] | -1.12[-5.7;4.34] |
| **Mortality Risk Score** | **MTE** | **NDE** | **NIE** | **PM** |
| Father's occupational position | 0.86[-2.30;3.09] | 0.76[-2.44;2.96] | 0.08[-0.14;0.17] | 0.13[-0.99;0.56] |
| Financial conditions in childhood | -1.25[-5.75;1.17] | -1.4[-5.82;1.07] | 0.07[-0.04;0.21] | -0.03[-1.48;1.44] |
| Father's education | 0.65[-2.67;3.59] | 0.53[-2.80;3.52] | 0.03[-0.11;0.2] | 0.06[-0.81;1.06] |
| Mother's education | 0.31[-5.72;1.93] | 0.17[-5.93;1.83] | 0.12[-0.02;0.18] | 0.43[-1.25;1.73] |

SE, Socioeconomic; MTE, Marginal Total Effect; NDE, Natural Direct Effect; NIE, Natural Indirect Effect; PM, Proportion Mediated
Counterfactual mediation analysis for the association between early-life SE indicators (exposure), SE indicators and lifestyle factors in adulthood (en-bloc mediators: smoking, alcohol intake, sedentary behavior, BMI, adulthood SE indicators), and markers of epigenetic aging. All models included age, sex, and study center as fixed-effect confounders, and familial structure as random-effect confounder.

We further show the counterfactual mediation analyses for the association between socioeconomic factors in adulthood, lifestyle factors, and markers of epigenetic aging in **Table 3**. We found that the associations between three socioeconomic indicators (household income, experiencing financial difficulties, and forgoing healthcare due to economic reasons) and DunedinPoAm38 and GrimAge EAA were jointly mediated by smoking, alcohol consumption, sedentary behavior, and BMI, with PM estimates ranging from 49% to 89%. Further, out of four socioeconomic indicators associated with the MS score, only associations involving occupational position were jointly mediated by lifestyle factors (own occupation: PM=31% 95%CI[6%;105%]).

**Table 3:** Counterfactual mediation estimates for the associations between SE indicators in adulthood, lifestyle factors, and markers of epigenetic aging (DAG: Figure 1B)

| **DNAmpHenoAge acceleration (Levine)** | **MTE** | **NDE** | **NIE** | **PM** |
| --- | --- | --- | --- | --- |
| Education | 0.44[-0.77;1.90] | 0.04[-1.21;1.55] | 0.00[-0.27;0.41] | -0.01[-2.78;2.27] |
| Occupational position | -0.07[-1.47;1.31] | -0.43[-1.81;0.96] | 0.23[-0.02;0.57] | -3.31[-6.69;4.88] |
| Household income | 1.01[-0.38;2.49] | 0.83[-0.62;2.43] | 0.06[-0.48;0.67] | 0.05[-1.90;1.80] |
| Financial difficulties | 1.16[-0.26;2.39] | 0.45[-0.94;1.76] | 0.25[-0.5;0.89] | 0.21[-1.74;3.09] |
| Forgoing healthcare due to fin. diff. | 0.39[-1.06;1.86] | 0.06[-1.39;1.49] | 0.40[-0.26;1.27] | 1.02[-4.25;11.82] |
| **DunedinPoAm38 age acceleration** | **MTE** | **NDE** | **NIE** | **PM** |
| Education | 0.01[-0.01;0.03] | 0.00[-0.02;0.01] | 0.01[-0.01;0.02] | 1.41[-8.66;9.77] |
| Occupational position | 0.00[-0.01;0.02] | 0.00 [-0.02;0.02] | **0.01[0.00;0.02]** | 4.53[-14.14;16.98] |
| Household income | **0.02[0.00;0.04]** | 0.01[-0.01;0.02] | **0.01[0.00;0.03]** | **0.75[0.13;2.49]** |
| Financial difficulties | **0.03[0.01;0.05]** | 0.01[-0.01;0.03] | **0.02[0.01;0.04]** | **0.71[0.39;1.45]** |
| Forgoing healthcare due to fin. diff. | **0.03[0.01;0.05]** | 0.01[-0.01;0.03] | **0.01[0.00;0.03]** | **0.49[0.12;1.6]** |
| **GrimAge acceleration** | **MTE** | **NDE** | **NIE** | **PM** |
| Education | 0.74[-0.21;1.63] | 0.24[-0.59;1.06] | 0.62[-0.18;1.26] | 0.83[-2.17;3.26] |
| Occupational position | 0.64[-0.31;1.39] | 0.04[-0.77;0.9] | **0.79[0.16;1.14]** | 1.24[-5.9;9.46] |
| Household income | **1.19[0.10;2.01]** | 0.49[-0.38;1.3] | **1.06[0.14;1.66]** | **0.89[0.19;3.38]** |
| Financial difficulties | **1.65[0.66;2.64]** | 0.53[-0.41;1.54] | **1.36[0.67;2.14]** | **0.83[0.45;1.65]** |
| Forgoing healthcare due to fin. diff. | **1.53[0.19;2.78]** | 0.58[-0.51;1.63] | **1.08[0.20;2.11]** | **0.70[0.19;2.13]** |
| **Mortality Risk Score** | **MTE** | **NDE** | **NIE** | **PM** |
| Education | **0.18[0.03;0.37]** | 0.16[-0.03;0.34] | 0.04[-0.02;0.13] | 0.25[-0.16;1.36] |
| Occupational position | **0.24[0.06;0.45]** | **0.19[0.00;0.39]** | **0.07[0.02;0.14]** | **0.31[0.06;1.05]** |
| Household income | **0.34[0.16;0.52]** | **0.25[0.06;0.41]** | 0.06[-0.01;0.15] | 0.21[-0.01;0.59] |
| Financial difficulties | 0.07[-0.1;0.29] | -0.03[-0.22;0.16] | 0.11[-0.01;0.26] | 1.52[-6.66;9.46] |
| Forgoing healthcare due to fin. diff. | **0.27[0.02;0.47]** | 0.15[-0.06;0.34] | 0.09[-0.02;0.26] | 0.39[-0.21;1.52] |

SE, Socioeconomic; MTE, Marginal Total Effect; NDE, Natural Direct Effect; NIE, Natural Indirect Effect; PM, Proportion Mediated
Counterfactual mediation analysis for the association between adulthood indicators (exposure), lifestyle factors (en-bloc mediators: smoking, alcohol intake, sedentary behavior, BMI), and markers of epigenetic aging. All models included age, sex, early-life SE indicators and study center as fixed-effect confounders, and familial structure as random-effect confounder

### 3.4 Supplementary analyses

#### 3.4.1 Intrinsic markers of epigenetic aging

Further, we estimated *intrinsic* markers of epigenetic aging by accounting for WBC composition using the Houseman method, but found no major differences when compared to the main analyses (**Supplementary Figure 2**).

#### 3.4.2 Stratification by age

Finally, we repeated the main analyses by stratifying the associations between socioeconomic indicators and markers of epigenetic aging at 65 years of age (**Supplementary Figures 4-5**). Overall, we observed similar associations in individuals aged <65years (**Supplementary Figure 3**) and the complete study sample (Figures 1). In older individuals, we observed much weaker gradients for associations involving adulthood socioeconomic factors and markers of epigenetic aging, and no clear associations for childhood socioeconomic factors (**Supplementary Figure 4**).

## 4 Discussion

In this population-based study, we found that socioeconomic disadvantage, whether experienced in childhood or adulthood, was consistently associated with biological age acceleration, as indexed using markers of epigenetic aging. Overall, we observed that the obtained signals were stronger for financial conditions in childhood, as well as household income and experiencing financial difficulties in adulthood than for other socioeconomic measures. Additionally examining life-course socioeconomic trajectories, we concluded that individuals experiencing adversity in childhood *and* adulthood displayed detrimental epigenetic aging patterns, implying “scarring effects” which result from early-life privation, along with more “proximal” physiological consequences resulting from financial hardship experienced in everyday life. Finally, by investigating the intermediate mechanisms through which socioeconomic factors operate, we found strong mediating effects by lifestyle factors for the association between socioeconomic indicators in adulthood and markers of epigenetic aging.

Using childhood socioeconomic indicators, we found that adverse financial conditions in childhood and a low father’s occupation were associated with increased Levine EAA, GrimAge EAA, and MS score. These results are in line with previous investigations, which reported that early-life socioeconomic disadvantage was related to increased epigenetic age acceleration, pointing to the importance of early-life material and financial circumstances in shaping the epigenetic signature [32, 41]. From a biological perspective, these findings may be related to the fact that epigenetic aging occurs faster in early-life, which may therefore represent a “critical” or “sensitive” period, when the body is more susceptible to adverse external exposures [24, 32]. To specifically determine whether childhood socioeconomic circumstances affect epigenetic aging via exposures in adulthood (“chains of risk model”) or by leaving a biological imprint in early-life (“critical period model”), we conducted a counterfactual mediation analysis using socioeconomic and lifestyle factors in adulthood as mediators. We found modest *marginal total effects* for the association between linearized childhood socioeconomic factors and markers of epigenetic aging, and no evidence for a mediating effect by exposures in adulthood [24]. Although some of the obtained results may support the “critical” period model (null *indirect* effect), the exact life-course mechanisms cannot be inferred based on these analyses only, given the absence of a consistent *marginal total effect* between linearized childhood socioeconomic indicators (main exposure) and markers of epigenetic aging (main outcome) in this particular framework (please see *Strengths and limitations*) [24, 34].

Further examining the associations between socioeconomic indicators in adulthood and markers of epigenetic aging, we found that a low occupational position, a low household income, experiencing financial difficulties in everyday life, and forgoing healthcare due to economic reasons were associated with detrimental epigenetic aging patterns, independently of the effect of childhood socioeconomic circumstances. These findings complement the growing body of evidence for an association between adverse socioeconomic conditions in adulthood and epigenetic age acceleration, reporting that constant financial pressure, poor living conditions, and economic hardship lead to chronic stress, eventually resulting in biological weathering and premature aging [42, 43]. Using counterfactual mediation models, we found that an important proportion of the association between adulthood socioeconomic factors and markers of epigenetic aging was substantially mediated by health behaviors and BMI in adulthood. These findings are in line with previous research reporting a strong mediating effect by lifestyle-related factors, including smoking, alcohol consumption, and obesity, which display a strong social patterning, and were previously associated with aberrant epigenetic methylation patterns [17, 28, 44, 45].

To gain insight into the life-course mechanisms underlying the association between socioeconomic factors and epigenetic aging, we further included life-course socioeconomic trajectories as an additional exposure variable, whereby a stable low trajectory (low father’s occupation in childhood and low own occupation in adulthood) was consistently associated with the highest levels of epigenetic age acceleration and mortality risk score. The joint analysis of childhood, adulthood, and life-course socioeconomic exposures thus showed that being *ever* exposed to socioeconomic adversity leads to detrimental epigenetic aging patterns, with early-life socioeconomic circumstances leaving a “scarring effect”, whilst subsequent, financial and economic hardship in adulthood affect epigenetic aging through other mechanisms [24]. From a life-course perspective, these results tend to point out towards the “sensitive period” effect model, a type of “chains of risk” model, implying that individuals experience periods or time-windows when the body is *more* sensitive to adverse external exposures than it would be at other times, although the observed physiological consequences may be influenced or modified by subsequent exposures [24].

Comparing the socioeconomic gradients across markers of epigenetic aging, we further observed that indicators of financial hardship in childhood and adulthood were consistently associated with mitigated epigenetic aging. These results suggest that all four epigenetic markers capture the same dimensions of socioeconomic status, namely financial and material deprivation across the life-course, despite the fact that the included aging markers have been developed using different training data and methodologies [16, 20, 22, 28].

Finally, when stratifying the associations between socioeconomic factors and markers of epigenetic aging by chronological age, we found similar associations in younger individuals (<65 years) and the full study set, while there were larger CIs for associations involving adulthood socioeconomic indicators and epigenetic markers in older individuals, and no clear associations involving childhood socioeconomic indicators. These results may be related to the much smaller sample size and lower statistical power in the “older” group (N _<65y_ = 509, N ≥65y = 175), but also to survival bias, as socioeconomically disadvantaged individuals “age” and die earlier, thus being less numerous in the “older” group [32, 46]. Furthermore, other plausible explanations may include age-related confounding factors, such as medication intake or menopausal status, which were previously associated with epigenetic age acceleration [47, 48].

### 4.1 Strengths and limitations

Our study has several strengths, the first being the use of multiple socioeconomic indicators in childhood, adulthood, and across the life-course. This has allowed us to identify which dimensions of socioeconomic status were the most consistently related to markers of epigenetic aging: in this particular case, financial and material hardship in childhood and adulthood, likely operating according to the “sensitive period” life-course model. Second, we compared four second generation markers of epigenetic aging, further highlighting the detrimental role of financial difficulties in childhood and adulthood for all included markers. Finally, this is one of the few studies formally quantifying the joint contribution of intermediate factors to the association between life-course socioeconomic indicators and markers of epigenetic aging.

Our study also has important limitations to acknowledge. First, the relatively small sample size has likely resulted in limited statistical power, which restricts the ability to detect smaller effect-size associations, particularly the ones between certain early-life socioeconomic factors and markers of epigenetic aging. Second, the reporting of early-life/childhood socioeconomic circumstances may be subject to recall bias, which may further “dilute” associations involving these indicators [49]. Third, in order to implement counterfactual mediation analysis, we assumed a linear (or dose-response) effect of the main exposure variables (childhood/adulthood socioeconomic factors) on markers of epigenetic aging, which may have led to biased associations, precluding the identification of specific life-course models. Finally, two out of four markers of epigenetic aging were computed using incomplete sets of CpGs, including GrimAge (947/1030 CpGs), and MS score (8/10 CpGs), which may have resulted in a suboptimal calculation of these scores.

### 4.2 Conclusion

In summary, our findings provide further evidence for an inverse gradient between socioeconomic circumstances across the life-course and detrimental epigenetic aging patterns, with an emphasis on intermediate and life-course mechanisms underlying this relation. Future research shall focus on further determining which socially-driven mechanisms lead to accelerated epigenetic aging, particularly focusing on the role of chronic stress, but also quantifying the overall contribution of altered epigenetic signals in the occurrence of diseases and mortality.

## Data Availability

All data produced in the present study are available upon reasonable request to the authors

## 5 Acknowledgments and funding

The authors would like to express their gratitude to Dr. Ake Lu and Prof. Steve Horvath (UCLA – Department of Human Genetics) for providing precious assistance for the calculation of the GrimAge epigenetic clock.

This work was supported by the Lifepath project, which is funded by the European commission (Horizon 2020 grant 633666), the Swiss state secretariat for education, research and innovation – SERI, the Swiss National Science Foundation, the Medical Research Council and the Portuguese Foundation for Science. Silvia Stringhini was supported by the Ambizione PROSPER grant (Grant PZ00P3_147998) from the Swiss National Science Foundation. Dusan Petrovic was supported by the Doc.Mobility grant (Grant P1LAP3_178061) from the Swiss National Science Foundation. The SKIPOGH study is funded by a grant from the Swiss National Science Foundation (Grant 140331) and by intramural support of Lausanne, Geneva, and Bern University Hospitals. The funders hand no role in study design, data collection and analysis, manuscript writing, and in the decision to submit.

## 7 Data and code availability

SKIPOGH data is not public, but may be made available upon a formal request submitted to the SKIPOGH steering board. For further details about the study design and the phenotypes available in SKIPOGH, please see https://www.maelstrom-research.org/study/skipogh.

The computer code (R software scripts) may be made available upon a request submitted to Dusan Petrovic.

## 8 Ethics statement

The SKIPOGH study was approved by relevant local or national ethics committees and all procedures performed in these studies were in accordance with the 1964 Helsinki declaration and its later amendments or comparable ethical standards. All participants gave written informed consent. This work does not contain any studies with animals performed by any of the authors.

## 9 Author contributions

**Dusan Petrovic**: formal analysis, investigation, methodology, writing (original draft and review/editing). **Cristian Carmeli**: conceptualization, data curation (processing), formal analysis, investigation, methodology, supervision, writing (review & editing). **José Luis Sandoval**: investigation, methodology, writing (review & editing). **Barbara Bodinier**: methodology, software, writing (review & editing). **Marc Chadeau-Hyam**: methodology, software, writing (review & editing). **Stephanie Schrempft**: investigation, writing (review & editing). **Georg Ehret**: data curation, investigation, writing (review & editing). **Nasser Abdalla Dhayat**: data curation, investigation, writing (review & editing). **Belén Ponte**: data curation, investigation, writing (review & editing). **Menno Pruijm**: data curation, investigation, writing (review & editing). **Emmanouil Dermitzakis**: data curation, investigation, writing (review & editing). **Paolo Vineis**: project administration, funding acquisition, data curation, investigation, writing (review & editing). **Sémira Gonseth-Nusslé**: data curation, investigation, writing (review & editing), methodology. **Idris Guessous**: data curation, investigation, writing (review & editing). **Cathal McCrory**: investigation, methodology, supervision, writing (review & editing), **Murielle Bochud**: project administration, funding acquisition, conceptualization, data curation, methodology, supervision, writing (review & editing), **Silvia Stringhini**: conceptualization, project administration, funding acquisition, data curation, methodology, investigation, supervision, writing (original draft, review & editing).

## 10 List of abbreviations

BMI: Body mass index
CI: Confidence interval
CpG: Cytosine-phospho-Guanine dinucleotide
DAG: Directed acyclic graph
DNA: Deoxyribonucleic acid
DNAmPhenoAge: Second generation *phenotypic* DNA methylation epigenetic clock (Levine)
DunedinPoAm: Second generation Dunedin Pace of Aging Methylation epigenetic clock
EAA: Epigenetic age acceleration
GrimAge: Second generation GrimAge epigenetic clock
MS: Mortality risk score
MTE: Marginal total effect
NDE: Natural direct effect
NIE: Natural indirect effect
OR: Odds ratio
PM: Proportion mediated
SD: Standard deviation
SE: Socioeconomic (factors)
SKIPOGH: Swiss Kidney Project on Genes in Hypertension
WBC: White blood cells

---

**Supplementary Figure 1:**
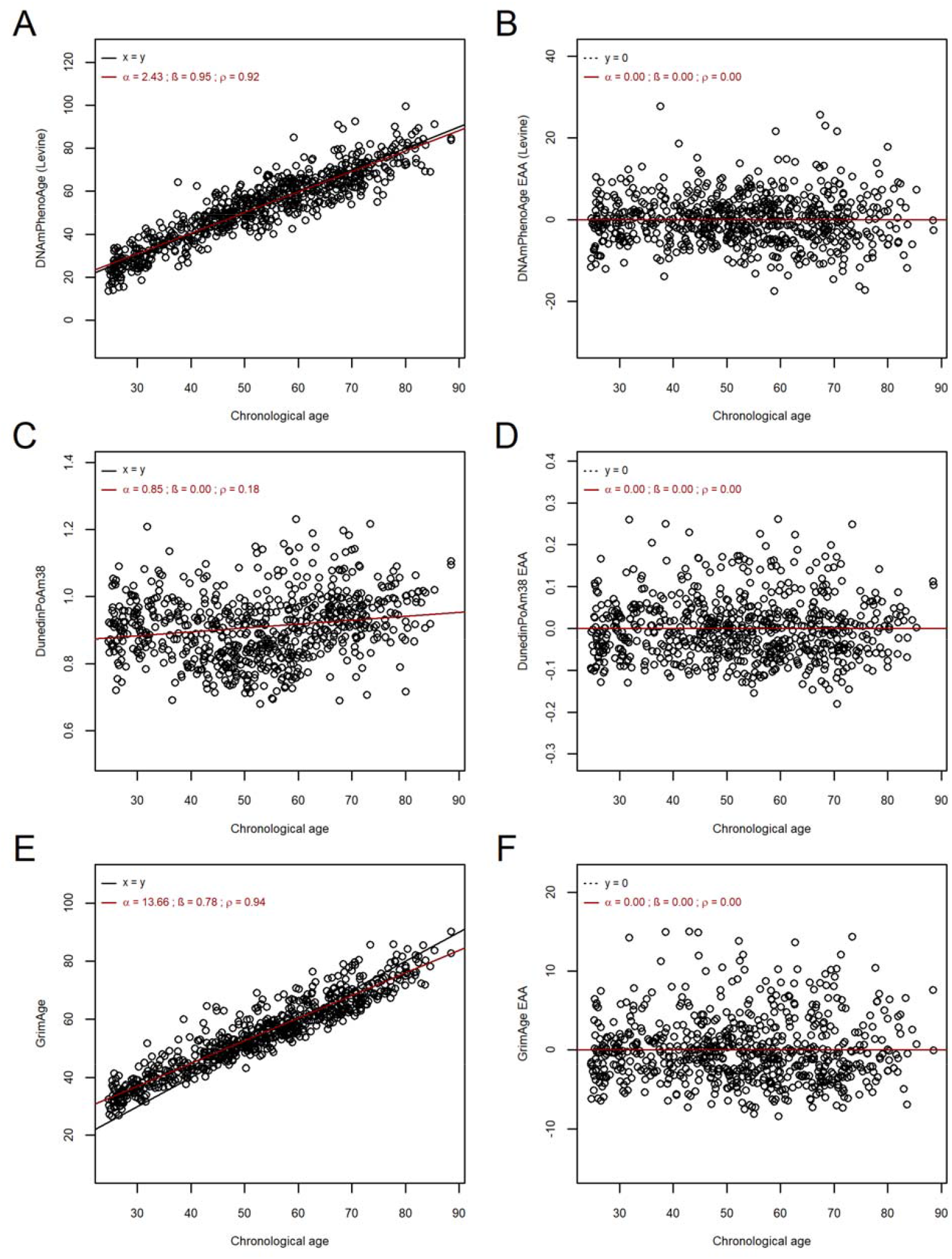
Scatterplots depicting linear associations between chronological age and second generation Levine, GrimAge, and DunedinPoAm38 epigenetic clocks (ACE), and between chronological age and epigenetic age acceleration (BDF) α: Intercept; β: Slope; ρ: Correlation coefficient ; EAA, Epigenetic age acceleration

**Supplementary Figure 2:**
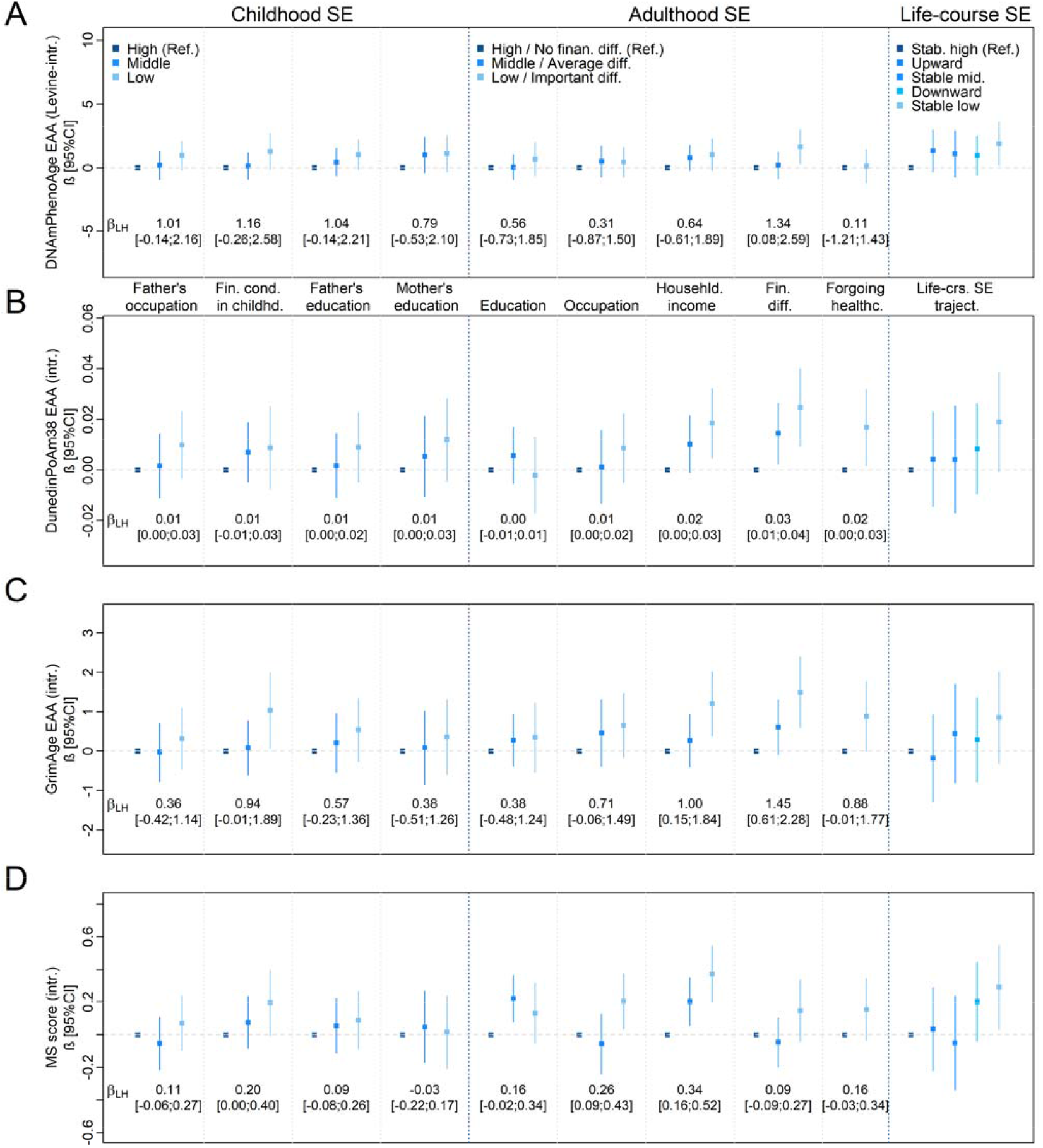
Regression coefficients for the associations between life-course socioeconomic factors and *intrinsic* markers of epigenetic aging EAA, Epigenetic age acceleration; Fin. cond., financial conditions; Fin. diff., Financial difficulties; Intr., Intrinsic; Life-crs. SE traject, Life-course socioeconomic trajectories; LH, lowest versus highest; SE, socioeconomic indicators Forgoing healthcare due to economic reasons was used as a two-categorical predictor: Not forgoing healthcare (Ref.); Forgoing healthcare Regression models for the association between life-course socioeconomic indicators (categorical predictor) and second generation *intrinsic* epigenetic age acceleration (ABC, continuous outcome) and mortality risk score (D, discrete outcome), adjusted for sex, age, center, season, familial structure (random effect). Associations involving socioeconomic indicators in adulthood were additionally adjusted for early-life socioeconomic indicators (father’s occupation, financial conditions in childhood, father’s education, and mother’s education) β_LH_: Regression coefficient for socioeconomic indicators used as *continuous (linearized)* predictors (Lowest vs. Highest) Missing data was imputed for socioeconomic indicators (childhood, adulthood, life-course) using multivariate imputation by chained equations and by hypothesizing missingness at random (5 imputed datasets, 50 iterations)

**Supplementary Figure 3:**
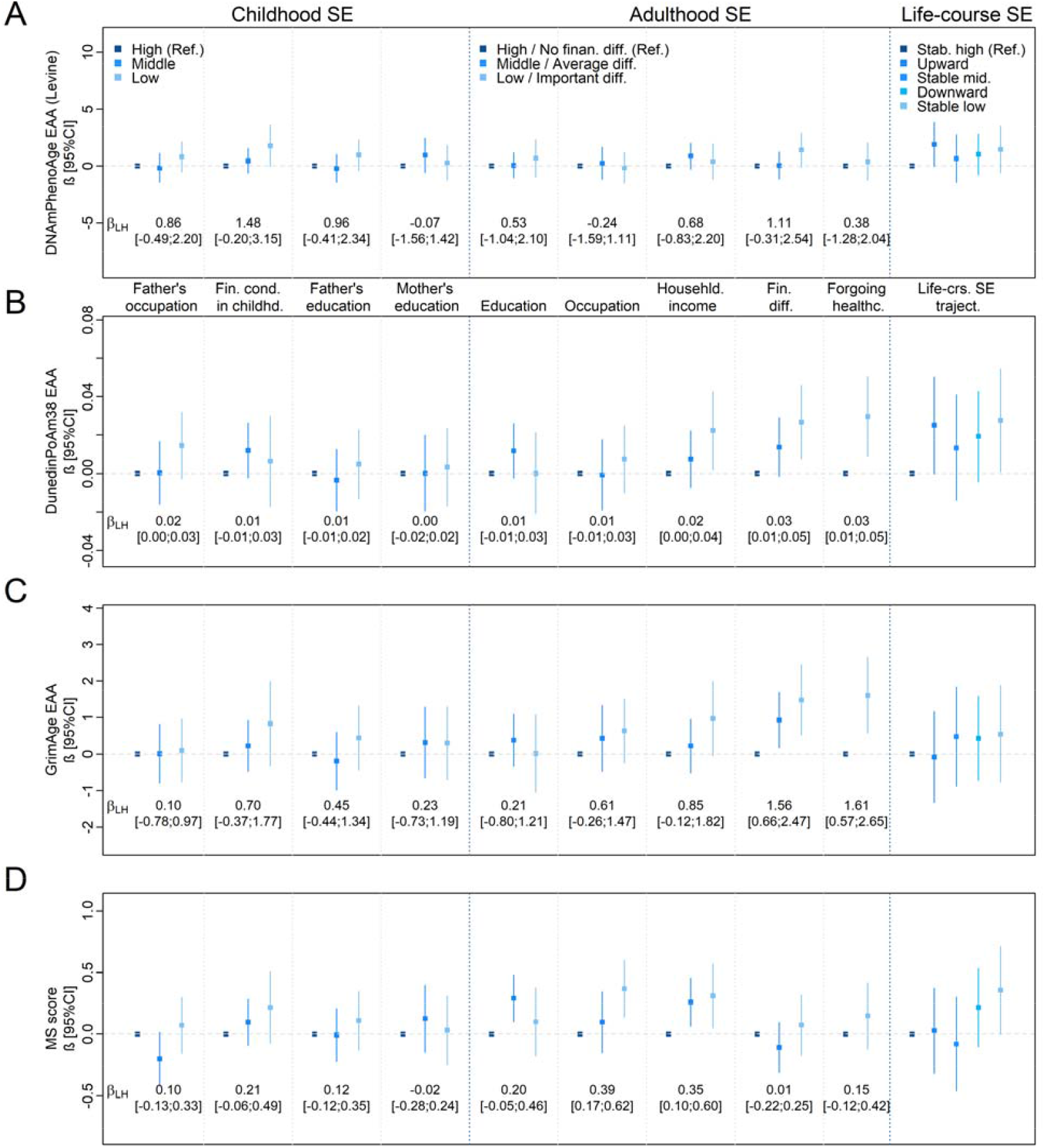
Regression coefficients for the associations between life-course socioeconomic factors and markers of epigenetic aging in individuals aged <65 years (N=509) EAA, Epigenetic age acceleration; Fin. cond., financial conditions; Fin. diff., Financial difficulties; Life-crs. SE traject, Life-course socioeconomic trajectories; LH, lowest versus highest; SE, socioeconomic indicators Forgoing healthcare due to economic reasons was used as a two-categorical predictor: Not forgoing healthcare (Ref.); Forgoing healthcare Regression model for the association between life-course socioeconomic indicators (categorical predictor) and second generation epigenetic age acceleration (ABC, continuous outcome) and mortality risk score (D, discrete outcome), adjusted for sex, age, center, season, familial structure (random effect). Associations involving socioeconomic indicators in adulthood were additionally adjusted for early-life socioeconomic indicators (father’s occupation, financial conditions in childhood, father’s education, and mother’s education) β_LH_: Regression coefficient for socioeconomic indicators used as *continuous (linearized)* predictors (Lowest vs. Highest) Missing data was imputed for socioeconomic indicators (childhood, adulthood, life-course) using multivariate imputation by chained equations and by hypothesizing missingness at random (5 imputed datasets, 50 iterations)

**Supplementary Figure 4:**
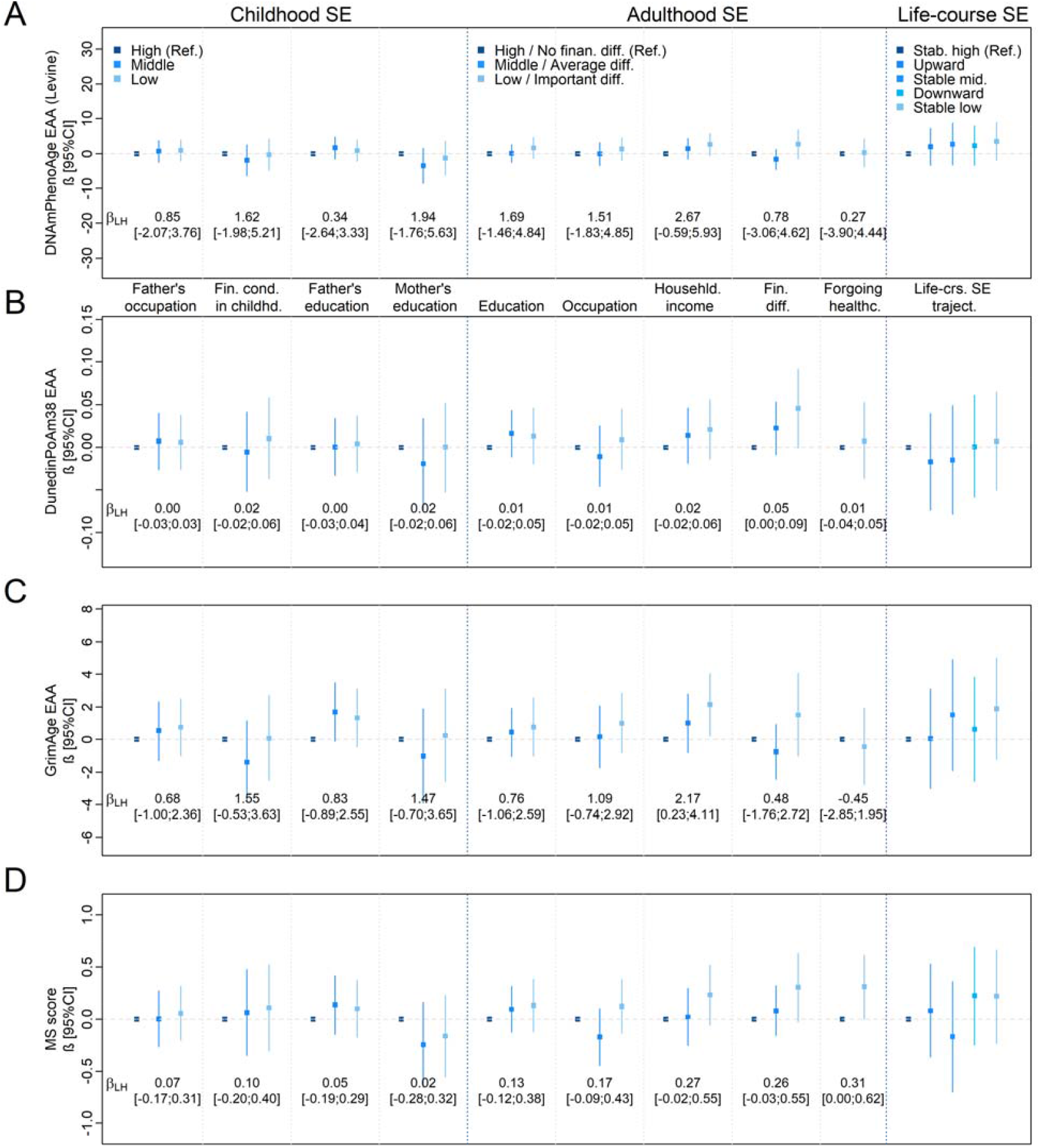
Regression coefficients for the associations between life-course socioeconomic factors and markers of epigenetic aging in individuals aged≥65 years (N=175) EAA, Epigenetic Age acceleration; Fin. cond., financial conditions; Fin. diff., Financial difficulties; Life-crs. SE traject, Life-course socioeconomic trajectories; LH, lowest versus highest; SE, socioeconomic indicators Forgoing healthcare due to economic reasons was used as a two-categorical predictor: Not forgoing healthcare (Ref.); Forgoing healthcare Regression model for the association between life-course socioeconomic indicators (categorical predictor) and second generation epigenetic age acceleration (ABC, continuous outcome) and mortality risk score (D, discrete outcome), adjusted for sex, age, center, season, familial structure (random effect). Associations involving socioeconomic indicators in adulthood were additionally adjusted early-life socioeconomic indicators (father’s occupation, financial conditions in childhood, father’s education, and mother’s education) β_LH_: Regression coefficient for socioeconomic indicators used as *continuous (linearized)* predictors (Lowest vs. Highest) Missing data was imputed for socioeconomic indicators (childhood, adulthood, life-course) using multivariate imputation by chained equations and by hypothesizing missingness at random (5 imputed datasets, 50 iterations)

## Appendix A

### Socioeconomic factors in childhood (self-reported)

Father’s occupational position (“*What was your father’s socio-professional category when you were a child?)* was self-reported using ten pre-defined categories and subdivided into three main groups: “High” (Reference group - most favorable: superior managers, liberal professions, CEO-directors, university professors), “Middle” (qualified non-manual workers, middle-level executives, self-employed (craftsman/trade)), “Low” (qualified manual and unqualified manual workers, farmers).

Father and mother’s education (*“What is the highest level of education that your father/mother achieved when you were a child?”)* were self-reported using ten predefined categories, and further classified into three groups: “High” (Reference group - most favorable: university education, superior education (+3 years education after graduating high school)), “Middle” (high school graduates, education preparing for a profession: apprenticeship), “Low” (mandatory education, trade school diploma, no diploma).

Material and financial condition in childhood inquired whether participants had the following items/activities when they were a child (*“What were the items/activities your family had/benefited from when you were a child?”)*: a car, a TV, a domestic worker, a dishwasher, a telephone, enough heat at home, participating to a social or cultural association, leaving home during annual vacation, home ownership. Owing ≥7 items was classified as “High” (Reference group - most favorable), 4-6 items was classified as “Middle”, and ≤3 items was classified as “Low”.

### Socioeconomic factors in adulthood and life-course trajectories (self-reported)

Own last known occupational position was inquired as *“What is your current/last occupation?”*, and subsequently classified using the European Socioeconomic Classification system as following [50]: “High” (Reference group - most favorable: superior managers, liberal professions, CEO-directors, university professors), “Middle” (lower level executives: teachers, qualified technicians, nurses), “Low” (qualified manual workers, sales-assistants, clerks and unqualified workers).

Own education was asked as *“What is the highest level of education that you have achieved so far?”* using the same predefined defined categories and groups as for father’s or mother’s education.

Financial difficulties inquired whether the participant would face difficulties paying food, rent, charges, insurance, and loans throughout the month (*“Are there times during the month when you are facing real financial to meet your needs (food, rent, bills, insurance, debt,…)?”*, and was coded as following: “No difficulties” (Reference group - most favorable: “This has never happened”), “Average difficulties” (“Not now, but this has happened in the past), “Important difficulties” (“This has happened in the recent past”).

Forgoing healthcare due to financial reasons was asked as *“During the previous 12 months, has it ever happened to you, your partner, or your children to forgo using healthcare services due to financial reasons?”*, and self-reported by answering “No” (Reference group - most favorable) or “Yes”. Socioeconomic trajectories across the life-course were classified as following : “Stable high” (Reference group: high father’s occupation and high own occupation), “Upward mobile” (low father’s occupation and middle/high own occupation, or middle father’s occupation and high own occupation), “Stable middle” (middle father’s occupation, middle own occupation), “Downward mobile” (high father’s occupation and middle/low own occupation, or middle father’s occupation and low own occupation), “Stable low” (low father’s occupation and low own occupation).

## Appendix B: Markers of epigenetic aging

Levine’s DNA methylation phenotypic age (DNAmPhenoAge) is a second generation clock which has been developed using CpGs related to both chronological age *and* ten health-related biomarkers [16]. In the present sample, all 513 CpGs required for Levine’s DNAmPhenoAge calculation were available.

Dunedin Pace of Aging Methylation (DunedinPoAm38) is a second generation epigenetic clock, calibrated on changes in aging based on 18 physiological parameters for a period of 12 years (26y-38y) [28, 44]. All 46 CpGs required for DunedinPoAm38 were available in SKIPOGH’s database.

GrimAge is a second generation epigenetic clock which incorporates multiple CpG sites related to plasma protein levels, smoking patterns, age, sex and mortality [22, 44]. GrimAge is computed based on 1030 CpG sites, out of which 947 were available in the SKIPOGH database.

The epigenetic mortality risk score (MS) is computed according to risk quartiles of ten health-related CpG sites, and has been previously related with disease occurrence, frailty, mortality but also Levine’s DNAmPhenoAge [17, 20]. Out of the ten required CpGs, eight markers were available in SKIPOGH’s methylation database and used to compute a discrete score, ranging from zero (most favorable - no CpGs in the risk quartile) to eight (least favorable – all eight CpGs in the risk quartile).

Epigenetic age acceleration (EAA) was computed for Horvath’s, Hannum’s, Levine’s, DunedinPoAm38, and GrimAge as the residuals from the regression of epigenetic clocks (response) by chronological age, and used as continuous outcome variable, whilst the MS score was untransformed and used as a discrete outcome variable.

## Appendix C: Reporting and grouping of health behaviors

Smoking status was categorized as non-current vs. current smoking, the former including previous smokers. Alcohol intake was measured using questions on the number of alcoholic drinks usually consumed within a week and categorized as hazardous drinking (>3 daily alcoholic drinks for men; >2 daily alcoholic drinks for women) versus non-hazardous drinking. Physical activity was reported on a scale from 1 to 10, 1 corresponding to a complete sedentary lifestyle and 10 corresponding to manual work combined with sports practice. Based on this scale, three categories were subsequently defined: “Low” (1–4), “Middle” (5), and “High” (6–10).

